# Hearing aid interventions to mitigate cognitive impairment: A critical review of the literature and meta-analysis

**DOI:** 10.1101/2025.03.25.25324637

**Authors:** Sumeet Thosar, Nikita U. Savant, Daniel A. Llano

**Affiliations:** Kansas City University; Carle Illinois College of Medicine; Department of Molecular and Integrative Physiology, University of Illinois at Urbana-Champaign; Beckman Institute for Advanced Science and Technology, Urbana, IL

## Abstract

Aging-related hearing loss (ARHL) is epidemiologically linked to the development of Alzheimer’s Disease (AD). The mechanisms underlying this relationship are not known and have important therapeutic implications. If ARHL is causally linked to the development of AD, then correction of hearing loss via hearing aids should mitigate cognitive impairments in AD and more aggressive campaigns to treat ARHL, which is widely undertreated, would be warranted. Here, we critically examine the literature involving the use of hearing aids to treat ARHL and examine the impact of hearing aids on cognition. Although many studies report beneficial effects of hearing aids on cognition, most of these studies have significant flaws in their experimental design, making it difficult to judge their outcomes. In our selection process, we prioritized randomized studies and those with blinded and placebo-controlled outcomes. We evaluated a total of 10 papers that met inclusion criteria. Within our literature review, we found two randomized placebo-controlled studies that examined the impact of hearing aids on cognition in cognitively-impaired or vulnerable older individuals with minimal risk of bias. Meta-analysis of these two studies did not yield a statistically significant benefit of hearing aid use after 6 or 12 months of use. We propose that the current literature on this topic currently lacks compelling evidence to demonstrate that hearing aid use directly benefits cognition or delays AD. We further recommend strategies for improving clinical trial design to bring greater clarity to this important issue.

## Introduction

Two increasingly common aging-related disorders that significantly impact quality of life are aging-related hearing loss (ARHL) and Alzheimer’s Disease (AD). Approximately 40% of adults over the age of 65 will develop ARHL as defined by an increase in pure-tone hearing threshold of at least 25 dB in the better-hearing ear^1^. In addition, approximately 10% of individuals in this age group will develop clinical signs of AD^2^. By 2050, the U.S. population in this age range is expected to reach 83 million^3^. Thus, as our population ages, the numbers of individuals suffering from these two disorders are expected to substantially increase.

It has been known for at least 30 years that the presence of ARHL increases the risk of developing dementia, based initially on non-prospective studies^4, 5^. More recently, longitudinal population studies have confirmed this relationship. For example, the Baltimore Longitudinal Study of Aging followed 639 initially cognitively normal individuals for 12 years and found that the hazard ratio for developing incident dementia was 1.89 for mild, 3.0 for moderate and 4.94 for severe hearing loss, after adjusting for shared risk factors for both ARHL and dementia such as age, sex, race, education, diabetes mellitus, smoking and hypertension^6^. This relationship between ARHL and the later development of dementia has been replicated several times and a meta-analysis of 10 studies revealed an overall hazard ratio of 1.58^7^. Thus, the epidemiological relationship between ARHL and AD is highly reproducible and seen across multiple populations. These data support the Lancet Commission’s reports stating that ARHL is the number one modifiable mid-life risk factor for the development of AD, with a population-attributable risk of 7%^8^.

The robust relationship between ARHL and AD raises several questions with significant therapeutic implications. For example, what is the mechanism underlying this relationship? Does ARHL cause or worsen AD pathology? Or are ARHL and AD linked by a common third, as yet unidentified, factor? If ARHL contributes to the underlying pathology that leads to AD, then therapeutic interventions to treat ARHL may mitigate the risk of developing AD. One important therapeutic intervention for ARHL is the use of hearing aids. Unfortunately, hearing aids are only used by a small fraction of the population that could potentially benefit from them^9, 10^. A longitudinal study from 1997 to 2023 found that only 26 states and 1 territory in the U.S. had private insurance mandates for hearing aids, with varying exception policies between companies that limit mandated coverage for adults with hearing loss^11^. If ARHL is causally linked to AD, then a stronger argument can be made to support public health measures to encourage greater adoption of this technology, and potentially other more aggressive interventions such as cochlear implants.

The clinical literature has not added substantial clarity about any potential mechanistic relationship between ARHL and AD. A recent study found that hearing loss and AD may be linked by serum lipidomic changes^12^, suggesting the presence of an underlying metabolic abnormality common to both disorders. Interestingly, hearing loss in established AD individuals is relatively mild, with a recent meta-analysis showing only a 3 dB difference in peripheral thresholds in AD vs. age-matched control participants^13^. In general, evidence that individuals with AD have greater central hearing loss has been more compelling than what has been seen with peripheral hearing (reviewed in^14^). Alternative hypotheses have been introduced such that AD may in fact be a risk factor for ARHL, despite the temporal differences between the clinical onset of these illnesses^15^. Given that AD has likely at least a decade of subclinical changes that occur prior to the onset of memory loss^16, 17^, it is possible underlying AD pathology may be present prior to ARHL and may in fact lead to ARHL^15^. Ultimately, human studies are challenging to interpret in terms of assessment of causality since it is not possible to induce hearing loss in humans and test for the development of AD.

Animal models are thus useful to attempt to answer questions related to causality since hearing loss can be induced and signs related to AD pathology can be examined in these animals. To date, many studies have been conducted that induce hearing loss in a rodent model and later found evidence for hippocampal dysfunction^18-20^, tau deposition and/or hyperphosphorylation^18, 21-23^ or increased amyloid beta production and/or deposition (^24, 25^, reviewed in ^26^). However, clear evidence for a progressive neurodegenerative phenotype induced by hearing loss has not yet been uncovered. Further, the clinical data linking ARHL and AD are derived from a gradual auditory deafferentation as seen in presbycusis, rather than the more typical laboratory manipulation which is to induce hearing loss with acoustic trauma, which leads to the acute and high expression of inflammatory mediators and other signs of trauma^27, 28^. The systemic inflammatory response after noise damage may itself be toxic to highly metabolically active regions such as the hippocampus^29, 30^. Thus, typical laboratory approaches to study hearing loss have had limited ability to answer the question about ARHL and AD causality.

If ARHL exacerbates AD pathology, whether through worsening the pathological cascade leading to typical AD-related pathology, or by inducing social isolation or by taking up cognitive resources needed to complete cognitive tasks, treatment of ARHL with hearing aids would be predicted to improve cognitive function. Recent studies and meta-analyses have been conducted and reported to show that hearing aid use has beneficial effects on cognition^8, 31^. However, many of the studies that have examined the relationship between hearing aid use and cognitive function have been retrospective analyses and/or been compromised by the lack of placebo or other appropriate controls. Thus, these studies do not provide a clear picture of the effect of hearing aids. Placebos can have clear and potent positive and negative effects across multiple neurological conditions, including AD^32-34^. Hearing aids provide a particularly potent placebo response given the constant presence of two devices worn on the head and the human contact needed to tune the devices^35^. Thus, it is important to provide a detailed analysis of the impact of hearing aid interventions in the context of the design of these studies. Here, we critically evaluate the literature establishing the potential benefit of hearing aid use on AD as well as aging-related cognitive impairment not yet classified as AD.

## Methods

Criteria for inclusion in the current analysis are that 1) a study was prospective and allocated participants with ARHL into hearing aid and control groups, 2) a study reported the impact of hearing aid use or nonuse on cognitive outcomes and reported statistical significance of each and 3) that participants in the study had a form of aging-related cognitive impairment (such as AD) or are at risk for such by virtue of being over 50 years of age. Pilot studies were not included if the full-form publication was included^36^. PubMed, Mendeley and Scopus using the keywords “hearing aid” and “cognition” were used to screen and identify studies to be used in this analysis. Final date of search = December 30, 2024.

The Cochrane Risk of Bias Tool 2 (RoB2^37^) was used to evaluate the potential risk of bias in each study. RoB2 provides a comprehensive assessment across multiple dimensions of prospective studies and is designed to be used for individual clinical trials. RoB2 uses five sets of signaling questions to assess risk of bias across five domains: randomization process, deviations from the intended interventions, missing outcome data, measurement of the outcome and selection of the reported result. For its use in the current manuscript, the downloadable Excel tool was used (downloaded from https://www.riskofbias.info/welcome/rob-2-0-tool/current-version-of-rob-2) and produced three possible outcomes per category (low risk, some concerns, high risk) and an overall outcome. Studies not employing a randomized design (or where such information was not provided), were automatically placed into the “high risk” category for randomization process and for overall score. Studies using open label interventions (but randomized design) were given at least “some concerns” for deviations from the intended interventions and measurement of the outcome. Studies that report less than 50% of starting participants in the final assessment (or where such information was not provided) were given at least “some concerns” for missing outcome data. Studies with greater than five cognitive outcome metrics were given at least “some concerns” for selection of the reported result. Any study with at least one high risk category was deemed high risk overall.

Once studies with an overall “low risk” RoB2 score were identified, a meta-analysis was done on the one cognitive outcome that was comparable between the two studies: a fluid cognition composite score from Searchfield et al.^38^ and Digit Symbol Test from Nguyen et al.^39^, a marker of processing speed which correlates with measures of fluid intelligence^40^. To conduct a quantitative analysis of the two studies that met low risk RoB2 criteria, the “meta-analysis” function of SPSS was used with Hedges’ g as the effect size measure. A random-effects model was chosen given the significant differences in study design, participant characteristics and outcome metrics. Data were taken directly from tables provided in the publications of mean baseline, 6 month and 12 month timepoints, standard deviations and numbers of participants. For Searchfield et al., the “Standard” approach was assumed to be the therapeutic intervention despite the authors’ expectations to the contrary, given the improved cognitive outcomes in this group.

## Results

After screening greater than 600 studies and removal of duplicates, 10 studies were identified that met inclusion criteria (see Figure 1). Below, we describe the studies in chronological order from when they were published. These studies are listed in Table 1 and Cochrane RoB2 determinations are in Table 2. Beyond the Cochrane RoB assessment, the main factors that we considered in assessing the utility of the outcomes were 1) were the groups randomized?, 2) were the participants blinded to the intervention?, 3) were the assessors blinded to the intervention?, 4) did the trial track adherence with hearing aids (which is critical for the interpretation of negative data)? and 5) were the assessments primarily obtained using an auditory-delivered scale? The latter would bias the study in favor of hearing aid use as amplification may diminish the cognitive workload needed to complete an assessment, which may not reflect global cognitive performance and potential for disease modification.

**Figure 1:**
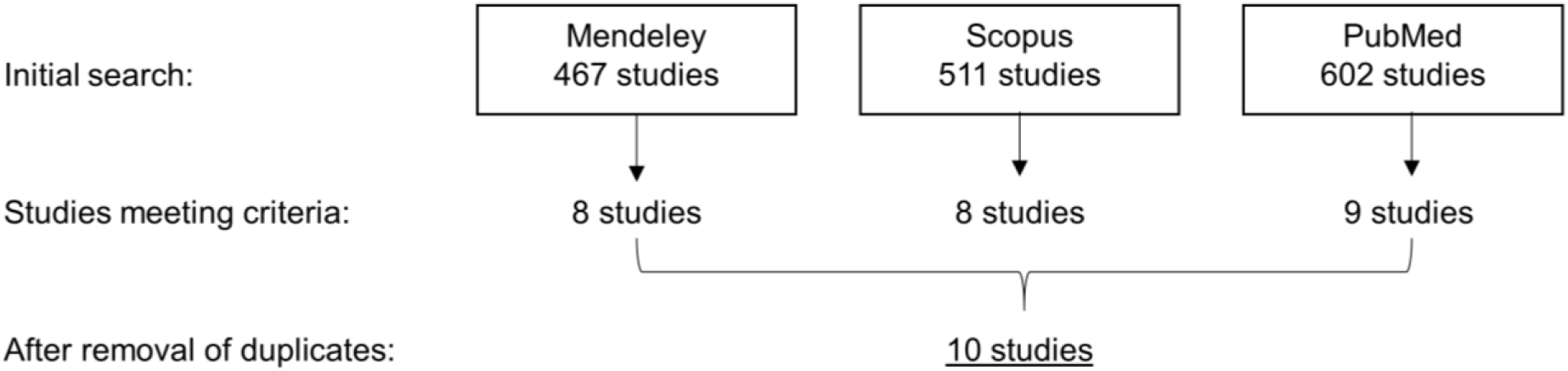
Search strategy for this study.

### Mulrow et al. 1990

The earliest study identified was Mulrow et al.^41^ from 1990. This study examined 194 participants over the age of 64 and randomly assigned them to a hearing aid group or waiting-list control group (Table 1). Waiting-list participants received their hearing aids at 4 months. Cognition was measured using the Short Portable Mental Status Questionnaire (SMPSQ). At 6 weeks, 85% of the active group reported at least 4 hours/day of use. At 4 months, hearing aid participants had statistically significantly better scores on the SMPSQ by 0.28 points (out of 10 points) than control participants. Note that at baseline, the hearing aid group had statistically significantly worse scores on the SMPSQ. Strengths of this study included its sample size and random allocation of groups. Weaknesses are that the study was unblinded and utilized a cognitive outcome measure that is entirely delivered verbally. The baseline differences in the SMPSQ are also concerning, given that the differences between groups represented regression towards the mean. On the RoB2 tool, this study was deemed high risk given the lack of blinding and the auditorily-derived outcome metrics (Table 2).

### Clemens-Tesch-Römer et al. 1997

Clemens Tesch-Römer^42^ in 1997 examined 112 aged cognitively normal aged participants with hearing impairment (Table 1). Active participants were fitted with a hearing aid and controls were those who self-selected to not wear a hearing aid for at least 6 months. These participants were found to have less hearing loss at baseline than active participants. Cognitive outcomes assessed at 6 months were Digit Symbol Substitution, Phonemic Fluency and Vocabulary testing. Hearing aid use was at least an average of 396 minutes (6.6 hours)/day. After 6 months of intervention, no differences were seen between the groups across cognitive outcomes. This study, which was non-randomized and non-blinded, was also confounded by differences in baseline hearing loss between experimental groups. The non-randomized and non-blinded nature of the study would have predicted an outcome in favor of hearing aids. It is not clear how to interpret the impact of worse baseline hearing loss on the effect of hearing aids on cognition since the relationship between degree of correction of hearing loss and cognition is also not known. On the RoB2 tool, this study was deemed high risk given the lack of randomization or blinding (Table 2).

### Van Hooren et al. 2005

In van Hooren et al.^43^, 122 participants aged 60 and older with mini-mental status exam (MMSE) score of at least 24 were enrolled to receive either 12 months of hearing aid use, or chose to not use hearing aids (Table 1). The latter group refrained from hearing aid use because of their less severe complaints or because of cost issues. Four out of 51 controls decided to use hearing aids during the study and were included in the control analysis based on intention to treat. A broad range of cognitive outcomes (Stroop Test, Concept Shifting Task, Letter-Digit Substitution Test, Visual Verbal Learning Test and Verbal Fluency) were assessed. At 12 months, the authors did not find a cognitive benefit of hearing aid use and found a decline in function in Stroop testing. In a subgroup of participants who wore the hearing aids for at least 8 hours/day, no benefits were seen (nor any worsening on Stroop Test). The deficits in study design here (lack of blinding, self-selected groups) as well as the factors in favor of the study design (size of the study, length of intervention) would have been expected to favor a positive outcome for hearing aid use on cognition. On the RoB2 tool, this study was deemed high risk given the lack of randomization or blinding and high number of cognitive outcome metrics assessed (Table 2).

### Choi et al. 2011

In Choi et al^44^, 29 participants were enrolled, where 18 were fitted with hearing aids and 11 were not. No information is provided about subject allocation or blinding. The average age of all participants was greater than 60 years. After 6 months, participants were tested on a computerized Korean visual verbal learning test (VVLT) whereby participants were shown 15 words and asked to recall the words. Thus, performance on this test was not fully dependent on auditory processing. Hearing aid participants showed significant improvements in word list recall and recognition at 6 months, whereas control participants did not. No information is given about hearing aid adherence. The lack of randomization and blinding (or at least information given about each of these), as well as the lack of information about adherence compromise the utility of this study. On the RoB2 tool, this study was deemed high risk given the lack of randomization or blinding, lack of information about the number of participants finishing the study and lack of information about adherence of hearing aid use (Table 2).

### Nguyen et al. 2017

In the Alzheimer Disease, Presbycusis and Hearing Aids (ADPHA) clinical trial^39^, Nguyen et al. conducted a multi-site 12 month study of the use of hearing aids in 48 participants over the age of 65 with AD (MMSE 10-28) on a common AD composite metric: the ADAS-Cog (Table 1). The ADAS-Cog is partially presented verbally (thus requiring auditory processing), but the secondary outcome measures of Free and Cued Recall and Digit Symbol Test were given via written instructions. Participants were randomized to receive either active or inactive hearing aids. After 6 months, all participants had active hearing aids. Only the hearing aid professional knew the activation status of the hearing aid – participants and study personnel remained blinded. Use for more than 5 hours/day was 83.3% in the active group. The finding that only 63.2% of the placebo group used their hearing aids for this duration suggests that functional unblinding may have been an issue, though this difference was noted not to be statistically significant. At 6 and 12 months post-fitting, no differences were seen between groups in any of the primary or secondary cognitive outcome measures. Overall, this study is deemed to be of high quality as it utilized a randomized, double-blinded format. If functional unblinding occurred (i.e., that the participants noticed whether their hearing aid was on), it would have been expected to enhance the treatment effect, which was not seen here. It is certainly possible that the intervention here was targeted too late in the pathological cascade (participants had an average MMSE score of approximately 19) for this intervention to be successful. This issue will be discussed in the Discussion section. On the RoB2 tool, this study was deemed low risk (Table 2).

### Karwani et al. 2018

Karawani et al^45^ studied 32 cognitively-normal individuals 60-84 years of age and randomly assigned 18 of them to receive hearing aids for 6 months (Table 1). The control group received hearing aids 6 months later. At 6 months, participants were tested with metrics taken from the National Institutes of Health Toolbox (www.nihtoolbox.org): List Sort Working Memory, Pattern Comparison Processing Speed Test and the Flanker Test. Participants and study staff were not blinded to assignment and participants were found to use hearing aids for 9.4 hours/day. At 6 months, working memory scores were found to improve in hearing aid users and to remain unchanged in control participants. There were no differences seen between the two groups on attention and speed of processing. It should be noted that the control group in this study was a “no-contact” group that had cognitive testing twice. The hearing aid group was seen four additional times for hearing aid assessments and evoked potential recordings, potentially inflating the placebo effect and facilitating comfort in the lab environment, as noted by the authors. More objective evoked potential metrics were also obtained, though the changes were primarily seen in the aided condition, suggesting that these differences may be in part related to acute increases in acoustic signal to noise rather than changes in brain physiology. On the RoB2 tool, this study was deemed some concerns given the lack of blinding (Table 2).

### Cuoco et al. 2021

Cuoco et al^46^ studied 56 participants over the age of 55 with hearing loss (Table 1). Hearing aid and control participants were self-selected and no mention of blinding is made. Multiple cognitive tests were performed at 6 months, including the Montreal Cognitive Assessment (MoCA), Digit Cancellation Test, Trails A, Rey Auditory Learning, Rey-Osterrieth Complex Figure Test, Clock Drawing Test as well as Raven’s Progressive Matrices. At 6 months, in the summary analysis, the hearing aid group showed improvements in Rey-Osterrieth Complex Figure Test and worsening on Ravens Progressive Matrices, though only the performance on the Ravens Progressive Matrices survived correction for multiple comparisons. Secondary analyses based on the severity of hearing loss were suggestive of cognitive improvements in the mild to moderate groups, but here the numbers of participants ranged from 4 to 6. No description of adherence is given. Similar to other studies, the lack of randomization, blinding and information about adherence compromise the utility of this study. On the RoB2 tool, this study was deemed high risk given the lack of randomization or blinding, lack of information about numbers of participants finishing the study, lack of information about hearing aid use and multiple cognitive outcome metrics assessed (Table 2).

### Lin et al. 2023

The Aging and Cognitive Health Evaluation in Elders (ACHIEVE) study^47^ is the largest of the studies meeting the criteria listed above. The ACHIEVE study recruited healthy older adults across multiple sites from the ongoing Atherosclerosis Risk in Communities study or de novo from the same sites. Participants had 30-70 dB of hearing loss and MMSE scores of 23 or above (25 if they had some college). These participants were randomized to either a three-year hearing aid intervention group or to receive successful aging health education. Participants were masked to the study hypothesis. A portion of the visits and assessments (from Mar 2020 to June 2021) were done over telephone due to COVID-19. The main outcome was a comprehensive neurocognitive battery containing Delayed Word Recall, Digit Symbol Substitution, Incidental Learning, Trail Making parts A and B, Logical Memory, Digit Span, Boston Naming, word fluency and animal naming as well as MMSE. These tests were adapted for administration via telephone during COVID shutdown. Thus, although the in-person battery contains a substantial number of tests that are not highly dependent upon auditory cues (e.g., trail-making), the telephone-modified version of this test presumably was entirely dependent upon auditory cues. Self-reported hearing aid use was high, with an average of 7.2 hours/day.

No differences were seen between groups on the primary outcome metric on the overall population (n=972). In the smaller subgroup recruited from the Atherosclerosis Risk in Communities group (n=238), a positive signal for hearing aid use in global cognition was seen, though the only subscale showing a significant difference was the language subscale, not executive function or memory. No differences were seen in either group in incident cognitive impairment. Although this study has been interpreted to support hearing aid use for overall cognitive improvement, its unblinded design and presumed reliance on auditory cues for some outcome assessments raise concerns. Additionally, the finding that only a subset representing 24% of the study population benefited from hearing aids, driven primarily by the domain most influenced by acoustic cues, calls into question the broader cognitive impact of such interventions. On the RoB2 tool, this study was deemed some concerns given the lack of blinding (Table 2).

### Sarant et al. 2024

In the ENHANCE study^48^, 262 participants aged 60 and older from two different cohorts were studied (Table 1). 160 participants were recruited from an audiology clinic (or clinics – there is no mention of the number of sites) to receive hearing aids. The hearing aid participants’ cognitive performance was compared to that of participants with hearing loss or normal hearing from the Australian Imaging, Biomarker and Lifestyle Flagship Study of Aging who did not use hearing aids. Outcomes were assessed using the Cogstate Brief Battery, a computerized assessment which is presented visually and contains a Detection test, Identification test, 1-Back test and Once Card Learning test. Hearing aid users showed good adherence (mean at different time points was 7.9-9.1 hours/day). Over the 36-month evaluation period, control participants showed declines in working memory, attention and psychomotor function, whereas the hearing aid group remained stable. No differences were seen in visual learning. No substantial change to the results was found when a subset of those without hearing loss was removed from the control group. It should be noted that despite the use of two independent cohorts, the groups were well-matched across most demographic variables and those for which matching was not adequate (e.g., education, physical activity) were adjusted for. Nevertheless, despite the comparatively adequate size and length of this study and attempts to control for demographic differences between active and control groups, the lack of randomization and blinding are significant impediments to the generalization of the findings. On the RoB2 tool, this study was deemed high risk given the lack of randomization or blinding, lack of information about numbers of participants finishing the study, lack of information about hearing aid use and multiple cognitive outcome metrics assessed (Table 2).

### Searchfield et al. 2024

The CogniAid trial by Searchfield et al.^38^ used a unique design compared to other studies. They screened participants aged 65 and older with hearing loss and enrolled 63 participants whose performance on the NIH toolbox composite score fell into the lower half of the screened population. All participants received hearing aids and were randomized to receive hearing aids fitted with “standard” nonlinear compressive approaches or “simple” linear amplification. Participants and raters were blinded to their allocation. Audiologists were unblinded. Participants were tested at baseline, 6 months and 12 months across 3 pre-specified domains of cognition: Fluid cognition composite score (Flanker, Dimensional Change Card Sort, Picture Sequence Memory, List Sorting and Pattern Comparison), Crystallized cognition composite score (Picture vocabulary and Reading test) and their average. Contrary to the author’s expectations, participants with standard fitting had significantly improve composite fluid intelligence scores compared to those with simple fitting and this difference grew at 12 months. No differences were seen on crystallized intelligence. Interestingly, no differences were seen Hearing Handicap Inventory for the Elderly or Modified Abbreviated Profile of Hearing Aid Benefit, though all groups showed benefits on these scales. Overall, this study design is seen as strong since participates were blinded and all participants received some degree of amplification, limiting the likelihood of functional unblinding. Unfortunately, the was no assessment of functional unblinding in this study. Circumstantial evidence for lack of unblinding is the similar high use of hearing aids across all groups (at least 7 hours per day on average). On the RoB2 tool, this study was deemed low risk (Table 2).

### Meta-analysis

Two out of the ten studies were deemed overall low risk of bias (Table 2,^38, 39^). They had one comparable cognitive outcome metric: fluid cognition composite score in the Searchfield et al. study and digit symbol test in the Nguyen et al. study. Digit symbol substitution is a metric of processing speed which has been shown to be correlated with fluid intelligence^40^. A total of 80 participants participated at the 6-month time point across both studies. Meta-analysis is not possible at 12-months because the Nguyen et al. study crossed over placebo participants (but not active participants) at 6 months. Combining these studies for the 6-month timepoint generated an overall effect size of 0.273 with 95% confidence interval of [-0.373, 0.919], with p=0.41, Figure 2.

**Figure 2:**
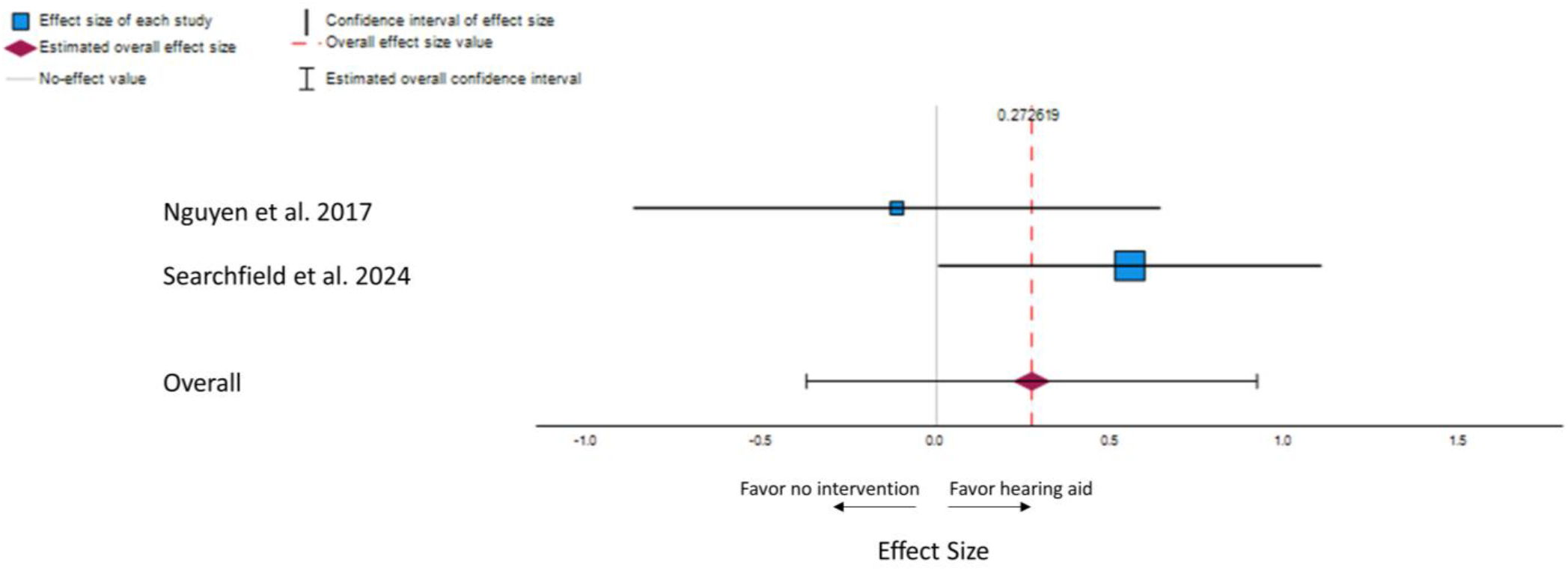
Forest plot of meta-analysis of data obtained after a 6-month intervention

## Discussion

In the current study, we examined 10 prospective studies on the use of hearing aids in aged populations and their impact on cognitive function. Six studies reported statistically significant favorable cognitive outcomes in participants receiving hearing aids. Five out of these six studies had a non-blinded interventions and outcomes, and thus would be considered “open-label.” Furthermore, several of these studies had non-randomized allocation or only showed cognitive benefit in a subgroup and/or in cognitive metrics fully dependent on auditory comprehension^41, 44, 45, 48^. Of the studies not showing a cognitive benefit of hearing aids, one used a randomized, double-blinded design^39^. This study was potentially compromised by enrolling participants that are possibly too far into the pathologic process of AD to benefit cognitively from most interventions. In this light, it is important to emphasize that older adults with cognitive impairment can benefit in terms of hearing performance from hearing aid use^49, 50^. Meta-analysis of the two studies deemed to be of low-risk did not yield a significant benefit of hearing aids for a marker of fluid intelligence.

Overall, the current analysis suggests that many of the benefits for cognition that have been described for hearing aids are not consistently seen across studies. In some cases, the benefits are likely driven by placebo effects, selection biases due to self-selection of active participants vs. controls, use of aided auditorily-delivered cognitive metrics and use of secondary subgroup analysis. Thus, these data suggest that the clinical literature does not yet unambiguously support the use of hearing aids as a means to diminish the risk of AD. We suggest that clinical equipoise still exists with respect to this question and that prospective, randomized, placebo-controlled trials are needed to determine the benefits of hearing aids for cognition. It should be emphasized that the data supporting hearing aids for improvement of speech intelligibility and quality of life have been consistently shown across multiple populations^51-55^, see^56^ for a meta-analysis on quality-of-life measures after hearing aid use. In this regard, the current study should only be interpreted in the context of hearing aids as a means to combat cognitive loss with aging and AD rather than a general commentary about the overall utility of hearing aids.

Previous meta-analyses and reviews have addressed whether hearing aids benefit cognition and have not all come to the same conclusion. For example, a recent meta-analysis of clinical studies concluded that the use of hearing aids led to improved cognitive outcomes in older adults^31^. However, most of studies included in this meta-analysis were retrospective analyses, did not measure cognition objectively and/or were observation studies of self-reported hearing aid use^57-66^ or did not have a control group^67, 68^, and were therefore not included in the current study. We suggest that the inclusion of non-randomized studies in this meta-analysis contributed to the positive conclusion in this analysis. Similarly, Sanders et al.^69^ and Yang et al.^70^ performed meta-analyses and included observational studies. Both of these meta-analyses generally found weak cognitive benefits that were isolated to the domain of executive function.

Retrospective and observational studies are heavily contaminated by placebo effects and self-selection (i.e., those with a favorable view of hearing aids are likely to actively seek them and report positive cognitive outcomes). It is important to recognize that although observation studies, particularly when providing results that are consistent with physiological principles, can be a powerful tool for hypothesis generation, they should not be used to infer causality. As an example, one of the main lessons from Women’s Health Initiative is that heavy reliance on observational studies can lead to erroneous conclusions and as such, clinical trials using randomized assignment, a double-blind design and the use of primary outcome measures should be the basis to determine if clinical interventions, such as hearing aids, lead to cognitive benefit^71^.

### Mechanisms underlying the ARHL-AD connection

Several mechanisms have been proposed that may explain the relationship between hearing loss and the development of dementia (reviewed in^26, 72^). For example, it is possible that diminished hearing function may lead to social isolation, loneliness, less engagement in stimulating activities, and consequently, an overall impoverished cognitive environment. Thus, individuals with hearing loss “underuse” their cognitive resources leading to a lesser cognitive function. Such diminishment in social engagement is a known risk factor for incident dementia^73^. It would be expected, then, that cognitive benefit would track with indicators of mood. Of the studies reviewed here that report a cognitive benefit of hearing aids, only Mulrow^41^ reported concomitant improvement in a depression rating scale, potentially consistent with this hypothesis, though subject-level data would be needed to confirm this relationship. Further work using parallel cognitive and mood metrics and explicit attempts to correlate them will help shed light on whether this relationship underlies the cognitive benefits of hearing aids described in the literature.

It has also been proposed that loss of hearing increases the cognitive workload of day-to-day life. In this scenario, successful listening requires engagement, greater working memory and selective attention, requiring activation of widespread areas of the cerebral cortex and diminishing the capacity for these regions to be used for other processes. This mechanism has elements that are at odds with the “impoverished input” mechanism outlined above, in that it provides a mechanism for widespread activation, rather than diminishment, of cortical circuits, both purportedly leading to cognitive dysfunction. In addition, although the argument that hearing loss leads to the utilization of cognitive resources may explain diminished online cognitive performance, it does not logically lead to structural damage associated with dementia, which has been noted to be associated with hearing loss^74-76^.

Hearing loss can also lead to plastic changes in the aging brain, such as downregulation of synaptic inhibition, increases in spontaneous activity, degradation of temporal processing, deviance detection and increases in markers of in the auditory system^77-83^. Such changes are thought to underlie many of the neurologic manifestations of hearing loss, such as tinnitus, hyperacusis and diminished speech perception^78, 84, 85^. Given the strong connections between auditory structures and other temporal lobe structures compromised in AD, it is possible that central auditory pathology could disrupt entorhinal and/or hippocampal circuits underlying AD. For any of these mechanisms, restoration of peripheral acoustic input would be expected to prevent later cognitive worsening – a result not apparent in the double-blinded clinical literature reviewed herein. Of course, it is likely that it would only be possible to reverse such changes with earlier interventions and that such gains would only be seen with larger studies.

Mechanisms that would be consistent with the lack of benefit of hearing restoration for cognition would be those where there existed a common pathology shared by both ARHL and AD, or if AD predisposed to ARHL. Although the latter mechanism appears inconsistent with the temporal sequence typically seen of ARHL followed by AD, as noted above, clinical AD is thought to be preceded by pathological changes by several years if not decades^16, 17^. Several studies have shown that amyloid-overexpressing mice have hearing loss^86-88^, suggestive of a reverse-causation mechanism. “Common-cause” mechanisms, such as metabolic, vascular or inflammatory changes, have also been suggested (reviewed in^89, 90^). Despite findings that the ARHL-AD association exists after adjusting for commonly-encountered metabolic pathologies such as diabetes, it does not exclude the possibility that other as yet unidentified copathologies may exist. Both the cochlea and hippocampus are noted to be highly metabolically vulnerable tissues that are subjected to aging-related oxidative stress. To maintain the endocochlear potential, the stria vascularis, inner/outer hair cells and supporting cells in the organ of Corti express high levels of markers of metabolic activity^91-93^. Indeed polymorphisms in genes involved in the regulation of oxidative stress significantly impact the onset age of ARHL^94^. Further, exposure to ototoxic stimuli such as loud noise increases markers of oxidative stress in the cochlea as well as in brain regions involved in AD such as the hippocampus^21, 29, 30^. Thus in our opinion, a common shared pathology may contribute to the ARHL-AD association.

### Limitations of current study

The main limitation of the current report is the small number of individual studies (n=2 studies) and participants (n=80 total participants across two studies) that qualified for inclusion into a formal meta-analysis. As mentioned above, previous attempts to combine studies into a meta-analysis included studies with a high risk of bias. Although inclusion of a large numbers of participants can add clarity to a meta-analysis by homogenizing variability across studies, including of studies with high risk of bias often drags outcomes of meta-analyses towards the biased outcomes of the largest studies^95^. An additional weakness is the inclusion of studies of two different populations into the meta-analysis: participants with AD in the Nguyen et al. study and those without a specific cognitive diagnosis in the Searchfield et al. study. It is possible that the lack of an effect in the Nguyen et al. study stems from the more advanced state of cognitive impairment in these participants – a state typically thought to be less reversible than preclinical stages. As such, combining these two studies may have led to an underestimate of the impact of hearing aids on cognition. Both limitations will be ameliorated as new, well-designed studies are introduced into the literature. In addition to the issues previously discussed, many of the studies not included in the meta-analysis also involved small sample sizes, which significantly limits the generalizability of the findings. Small participant numbers reduce the statistical power of the studies, making it more likely that non-significant results are due to insufficient power rather than a true lack of effect. Furthermore, as highlighted earlier, inconsistencies in the selection and measurement of clinical outcomes can obscure the potential impact of hearing interventions.

### Overall conclusions

Given the clinical equipoise that exists in the relationship between hearing aid use and cognition, further trials are needed. We propose that several criteria should be considered when designing an informative clinical trial to assess the benefits of hearing aids for the potential prevention of AD. First, allocation to hearing aids should be randomized and participants and assessors should be blinded to their treatment group. The ethics of random assignment given the quality-of-life benefits of hearing aids would need to be addressed proactively. Because of the lack of established disease-modifying capacity of hearing aids and the growing importance of the societal burden of AD, it can be argued that random assignment is an appropriate approach. If we take into consideration the compelling need to understand the effect of hearing aids on cognitive function alongside the logistical realities of hearing aid use, including the long delays between hearing aid candidacy and adoption of hearing aids (> 8 years,^96^), the high rates of hearing aid discontinuation (>30%^97^), especially in those with mild hearing loss, in whom the effect of hearing aids on safety is not as significant as for greater severities of loss, an argument can be made that there is negligible risk, and much to be gained by conducting a placebo-controlled trial^98^.

Blinding is particularly challenging in this sphere given that participants may correctly guess their group allocation (as has happened in previous attempts at placebo-controlled hearing aid trials in Brewster et al.^99^), but approaches to limit placebo effects, such as permitting activation of hearing aids in all participants but with differences in specific programming, as employed in Searchfield et al.^38^, may ensure that all participants would detect the presence of an active amplification device and thus would have roughly similar placebo effects. Further, this approach can help ensure that effects on ear canal resonance, which is affected by hearing aid insertion are similar. It is also important that outcomes be assessed in a non-aided manner to differentiate changes in brain state from the acute effects of amplification on cognitive performance. To determine the potential impact of hearing aids on mood and subsequent effects on cognition, measurement of mood and determination of its contribution to cognitive outcomes should be performed. To determine if hearing aid interventions have the potential to modify AD pathology, AD biomarkers, such as hippocampal volume or markers of amyloid beta and/or tau in serum, spinal fluid or brain should be employed. Serum markers are growing in their availability, can predict neural metrics and may make biomarker assessment particularly feasible for large-scale studies^100, 101^.

Given the heavy and growing burden of both ARHL and AD in our society, and current marketing approaches that make claims about the ability of hearing aids to delay dementia diagnoses^102^, it is imperative that the field move beyond observational studies and provide a solid basis by which we can understand this important relationship. Such knowledge will provide greater guidance to clinicians and patients about the importance of pursuing hearing aid fitting and will provide a greater understanding of the neurobiology of both ARHL and AD.

## Data Availability

All data produced in the present study are available upon reasonable request to the authors

## Acknowledgments

No grant funding was used to support this research. The authors thank Dr. Shraddha A. Shende, Department of Communication Sciences and Disorders, Illinois State University, Normal, IL, for her comments on the manuscript.

